# The LYMPH Trial - Comparing Microsurgical with Conservative Treatment of Chronic Breast Cancer Associated Lymphedema: Study Protocol of a Pragmatic Randomized International Multicentre Superiority Trial

**DOI:** 10.1101/2024.02.13.24302744

**Authors:** Elisabeth A Kappos, Yvonne Haas, Alexandra Schulz, Florian Peters, Shakuntala Savanthrapadian, Julia Stoffel, Maria Katapodi, Rosine Mucklow, Benedict Kaiser, Alexander Haumer, Stephanie Etter, Marco Cattaneo, Daniel Staub, Karin Ribi, Jane Shaw, Tristan M Handschin, Steffen Eisenhardt, Giuseppe Visconti, Gianluca Franceschini, Lorenzo Scardina, Benedetto Longo, Marcus Vetter, Khalil Zaman, Jan A Plock, Mario Scaglioni, Eduardo G González, Sergio D Quildrian, Gunther Felmerer, Babak J Mehrara, Jaume Masià, Gemma Pons, Daniel F Kalbermatten, Justin M Sacks, Martin Halle, Maximillian V Muntean, Erin M Taylor, Maria Mani, Florian J Jung, Pietro G di Summa, Efterpi Demiri, Dimitris Dionyssiou, Anne K Groth, Norbert Heine, Joshua Vorstenborsch, Kathryn V Isaac, Shan S Qiu, Patricia E Engels, Axelle Serre, Anna-Lena Eberhardt, Sonja Ebner, Matthias Schwenkglenks, Yvette Stoel, Cornelia Leo, Raymund E Horch, Phillip Blondeel, Björn Behr, Ulrich Kneser, Lukas Prantl, Daniel T Boll, Cristina Granziera, Lars G Hemkens, Nicole Lindenblatt, Martin Haug, Dirk J Schaefer, Christoph Hirche, Andrea L Pusic, Katrin Seidenstücker, Yves Harder, Walter P Weber

**Affiliations:** Department of Plastic, Reconstructive, Aesthetic and Hand Surgery, University Hospital Basel, Basel, Switzerland; University of Basel, Basel, Switzerland; Department of Clinical Research, University Hospital Basel, Basel, Switzerland; Department of Clinical Research, University of Basel, Founding Member Patient Advocacy Group Oncoplastic Breast Consortium, Basel, Switzerland; Patient Advocacy Group, Oncoplastic Breast Consortium, Basel, Switzerland; Department of Angiology, University Hospital Basel, Basel, Switzerland; Quality of Life Office, International Breast Cancer Study Group, a division of ETOP IBCSG Partners Foundation, Bern; Careum School of Health, part of the Kalaidos University of Applied Sciences, Zurich, Switzerland; Department of Plastic and Hand Surgery, University Hospital Freiburg, Freiburg, Germany; Department of Woman and Child Health and Public Health, Division of Plastic Surgery, Fondazione Policlinico Universitario “A Gemelli” IRCCS, Rome, Italy; Department of Woman, Child Health and Public Health, Division of Breast Surgery, Fondazione Policlinico Universitario “A Gemelli” IRCCS, Rome, Italy; Department of Plastic and Reconstructive Surgery, University of Rome Tor Vergata, Rome, Italy; Department of Oncology, Cantonal Hospital Baselland, Liestal, Switzerland; Department of Medical Oncology, Lausanne University Hospital, Lausanne, Switzerland; Department of Plastic Surgery and Hand Surgery; Cantonal Hospital Aarau, Aarau, Switzerland; Department of Hand- and Plastic Surgery, Cantonal Hospital Lucerne, Lucerne, Switzerland; Center for Plastic Surgery, Center for Breast Cancer Surgery, Pyramide am See Clinic, Zurich, Switzerland; Division of Oncoplastic Surgery, Oncologic Institute Instituto de Oncología Ángel H Roffo, Buenos Aires British Hospital, University of Buenos Aires, Buenos Aires, Argentina; Department of Trauma Surgery, Orthopedic Surgery and Plastic Surgery, University Medical Center Göttingen, Göttingen, Germany; Department of Plastic and Reconstructive Surgery, Memorial Sloan Kettering Cancer Center, New York, NY, USA; Department of Plastic Surgery, Hospital de la Santa Creu i Sant Pau Universitat Autònoma de Barcelona, Barcelona, Spain; Department of Plastic, Reconstructive, and Aesthetic Surgery, Geneva University Hospital, Geneva, Switzerland; Division of Plastic and Reconstructive Surgery, Washington University School of Medicine, Saint Louis, Missouri, USA; Department of Reconstructive Plastic Surgery, Karolinska University Hospital, Stockholm, Sweden; Department of Plastic Surgery, Institute of Oncology, University of Medicine and Pharmacy “Iuliu Hatieganu”, Cluj-Napoca, Romania; Division of Plastic and Reconstructive Surgery, Department of Surgery, Harvard Medical School, Brigham and Women’s Hospital, Boston, Massachusetts, USA; Department of Plastic and Maxillofacial Surgery, Uppsala University Hospital, Uppsala, Sweden; Department of Plastic and Hand Surgery, Cantonal Hospital of Winterthur, Winterthur, Switzerland; Department of Plastic and Hand Surgery, University Hospital of Vaudois (CHUV), Lausanne, Switzerland; Department of Plastic Surgery, Aristotle University of Thessaloniki, Thessaloniki, Greece; Department of Plastic Surgery, Hospital Erasato Gaertner, Curitiba, Brazil; Department of Plastic Surgery, Caritas Hospital St. Josef, Regensburg, Germany; Division of Plastic Surgery, McGill University, Montreal, Quebec, Canada; Division of Plastic Surgery, University of British Columbia, Vancouver, Canada; Department of Plastic, Reconstructive and Hand Surgery, Maastricht University Medical Center, Maastricht, the Netherlands; Department of Radiation Oncology, University Hospital Basel, Basel, Switzerland; Department of Obstetrics and Gynecology, Cantonal Hospital Baselland, Liestal, Switzerland; Health Economics Facility, Department of Public Health, University of Basel, Basel, Switzerland; Institute of Pharmaceutical Medicine (ECPM), University of Basel, Basel, Switzerland; Institute of Therapies and Rehabilitation, Division of Physiotherapy, Cantonal Hospital of Winterthur, Winterthur, Switzerland; Breast Center, Kantonsspital Baden, Baden, Switzerland; Department of Plastic and Hand Surgery, University Hospital of Erlangen, Erlangen, Germany; Department of Plastic and Reconstructive Surgery, Ghent University Hospital, Ghent, Belgium; Department of Plastic,- Reconstructive and Aesthetic Surgery, Kliniken Essen-Mitte (KEM), Essen, Germany; Department of Hand-, Plastic and Reconstructive Surgery, Burn Center, BG Trauma Center Ludwigshafen, University Hospital Heidelberg, Ludwigshafen, Germany; Department of Plastic Surgery, University Hospital Regensburg, Regensburg, Germany; Department of Radiology and Nuclear Medicine, University Hospital of Basel, Basel, Switzerland; Neurology Clinic and Policlinic, Departments of Head, Spine and Neuromedicine, Biomedicine and Clinical Research University Hospital Basel, Basel, Switzerland; Pragmatic Evidence Lab, Research Center for Clinical Neuroimmunology and Neuroscience Basel (RC2NB), University Hospital Basel and University of Basel, Basel, Switzerlandy; Meta-Research Innovation Center at Stanford (METRICS), Stanford University, Stanford CA, USA; Meta-Research Innovation Center Berlin (METRIC-B), Berlin Institute of Health, Berlin, Germany; Department of Plastic and Hand Surgery, University Hospital Zurich, Zurich, Switzerland; Breast Center, University Hospital Basel, Basel, Switzerland; Department of Plastic, Hand and Reconstructive Microsurgery, Hand Trauma and Replantation Centre, BG Unfallklinik Frankfurt am Main, Frankfurt am Main, Germany; Department of Plastic Surgery, Sana Hospitals Düsseldorf, Düsseldorf, Germany; Department of Plastic, Reconstructive and Aesthetic Surgery, Ospedale Regionale di Lugano, Ente Ospedaliero Cantonale (EOC), Lugano, Switzerland; Faculty of Biomedical Science, Università della Svizzera Italiana (USI), Lugano, Switzerland

## Abstract

**Introduction:** Up to one fifth of breast cancer survivors will develop chronic breast cancer-related lymphedema (BCRL). To date complex physical decongestion therapy (CDT) is the gold standard of treatment. However, it is mainly symptomatic and often ineffective in preventing BCRL progression. Lymphovenous anastomosis (LVA) and vascularized lymph node transfer (VLNT) are microsurgical techniques that aim to restore lymphatic drainage. This international randomized trial aims to evaluate advantages of microsurgical interventions plus CDT vs CDT alone for BCRL treatment.

**Methods and analysis:** The effectiveness of LVA and/or VLNT in combination with CDT, which may be combined with liposuction, versus CDT alone will be evaluated in routine practice across the globe. BCRL patients will be randomly allocated to either surgical or conservative therapy. The primary endpoint of this trial is the patient-reported quality of life (QoL) outcome “lymphedema-specific QoL”, which will be assessed 15 months after randomization. Secondary endpoints are further patient reported outcomes (PROs), arm volume measurements, economic evaluations, and imaging at different timepoints. A long-term follow-up will be conducted up to 10 years after randomization. A total of 280 patients will be recruited in over 20 sites worldwide.

**Ethics and dissemination:** This study will be conducted in compliance with the Declaration of Helsinki and the ICH-GCP E6 guideline. Ethical approval has been obtained by the lead Ethics Committee ‘Ethikkommission Nordwest- und Zentralschweiz‘ (2023–00733, 22.05.2023). Ethical approval from local authorities will be sought for all participating sites. Regardless of outcomes, the findings will be published in a peer-reviewed medical journal. Metadata detailing the dataset’s type, size and content will be made available, along with the full study protocol and case report forms, in public repositories in compliance with the FAIR principles.

**Trial registration:** The trial is registered at https://clinicaltrials.gov (ID: NCT05890677) and on the Swiss National Clinical Trials Portal (SNCTP, BASEC project-ID: 2023-00733) at https://kofam.ch/de. The date of first registration was 23.05.2023.

**Strengths and limitations of this study:**

- This is a pragmatic, randomized, international, multicentre, superiority trial, which has the potential to impact the clinical practice of therapy for patients with chronic BCRL.
- The pragmatic design will reflect clinical practice, thereby directly providing applicable results.
- A comprehensive long-term follow-up will be conducted, extending up to 10 years, to assess and analyze long-term outcomes.
- Patient advocates were intensely involved throughout the trial design.
- To date, no multicentric RCT has compared microsurgical techniques (LVA and VLNT) possibly combined with liposuction with CDT alone, thereby limiting patient’s access to available treatment options.
- The assessment of treatment quality (both conservative and surgical) at various sites is challenging due to potential variations resulting from the pragmatic design, which may influence the study’s outcomes.

## INTRODUCTION

Approximately one in eight women will be diagnosed with breast cancer during her lifetime.^(1)^ Roughly one in five breast cancer survivors will develop chronic breast cancer-related lymphedema (BCRL). ^(1–7)^ Lymphedema (LE) refers to the localized condition of tissue swelling, following anomalous lymphatic fluid retention in the stroma due to compromised lymphatic pathways.^(8, 9)^ It is defined as chronic if related signs and symptoms last longer than three months and affect one or more areas of the body.^(10, 11)^ BCRL is associated with pain in the shoulder and arm, consequently leading to limitations in shoulder mobility.^(1–7)^ The incidence of BCRL can range from 7% to 30% annually, depending on treatment modalities and patient-specific risk factors.^(12)^ The risk of developing chronic BCRL and its severity is associated with various factors, including the extent of breast/axillary surgery, adjuvant radiatio therapy, (neo-) adjuvant chemotherapy, the number of positive lymph nodes, treatment on the dominant arm and obesity.^(10, 13–21)^

LE can be characterized as one of the most underestimated and debilitating morbidities among the complications affecting breast cancer survivors.^(22–24)^ Beside the negative consequences for patient’s quality of life (QoL), LE further exerts a two-fold negative economic impact: Firstly, there is a direct impact on the healthcare system as patients with chronic BCRL put a relevant strain on healthcare resources due to the lifelong need for physiotherapy, compression garments, and treatment of infections. Secondly, there are indirect costs arising from lost productivity when affected patients spend prolonged times absent from work, have reduced working capacities or even enter early retirement due to disablement as a consequence of LE.^(25)^

Current treatment guidelines recommend conservative complex physical decongestion therapy (CDT), which includes special massage techniques, manual lymph drainage, local compression, physical exercise and meticulous skin care.^(26)^ However, this treatment is mainly symptomatic and therefore offers limited benefits.^(27)^ Using microsurgical procedures, surgeons can enhance the lymphatic system’s ability to transport lymphatic fluid. This process could ultimately drain excess lymphatic fluid congested in the tissues.^(28)^ Lymphovenous anastomosis (LVA) and vascularized lymph node transfer (VLNT) represent two surgical treatment options that hold the potential for the restoration of proper lymphatic drainage. LVA tries to accomplish this objective by establishing anastomotic connections between lymphatic vessels and veins ^(29–31)^. Conversely, VLNT involves the transplantation of functional lymph nodes to regions of the arm lacking lymph nodes or featuring compromised lymph node function. This process induces lymphangiogenesis in order to stimulate the development of lymphovenous and lympholymphatic pathways ^(32–35)^. Liposuction can be complemented in addition to both microsurgical procedures in order to reduce excess volume.^(28, 36)^

Several observational studies have examined the mode of action and efficacy of both LVA and VLNT for the treatment of chronic BCRL. A number of systematic reviews analyzing these surgical methods have indicated better patient outcomes and low complication rates.^(37–44)^

To date (as of January 2024), there are three registered RCTs on ClinicalTrials.gov, comparing surgical to conservative treatment. The first one started in January 2016 and compares LVA with CDT in 100 plannend participants with BCRL in Norway. The primary outcome is arm volume change.^(45)^ Another registered RCT, which started in January 2019, has a similar design, and is conducted nationally in two centers in the Netherlands with 120 plannend participants. Here, the primary outcome is QoL after 12 months of follow-up measured with Lymph-ICF questionnaire as the primary endpoint.^(46)^ In the interim analysis of 46 patients per group, the LVA group showed significant improvement in physical and mental function domains of the Lymph-ICF questionnaire after 6 months, but no statistical difference in the total Lymph-ICF score or limb volume reduction was observed in either group.^(47)^ 42% in the LVA group reduced or ceased using compression garments, compared to none in the CDT group. ^(47)^ The third registered RCT started in October 2021 with 64 plannend participants. The trial is also comparing LVA versus CDT in a monocentric setting ^(48)^, similarly to the trial in Norway, measuring volume changes as a primary outcome. The only published RCT on surgical LE treatment was monocentric and compared 18 patients who underwent VLNT followed by CDT to 18 patients who received only CDT. However, all of the patients presented with early stage BCRL, limiting generalizability of findings and their clinical applicability.^(49)^ The primary endpoint, limb volume reduction, showed a 57% decrease in the surgical group (VLNT) compared to 18% decrease in the control group (CDT). Furthermore, patients in the surgical group reported less pain, overall functional improvements and experienced significantly reduced episodes of infection.^(49)^ Nevertheless, a deficiency persists in the medical literature, as no multicentric randomized controlled trial (RCT) has prospectively compared the treatment results of these two surgical interventions in comparison to CDT as a stand-alone treatment. This trial aims to provide patients and clinicians with the scientific evidence to make better-informed decisions for or against surgical therapy of BCRL. In this regard, the trial will answer the question if lymphatic surgery combined with CDT provides better QoL than the current standard treatment with CDT alone in patients with BCRL (primary objective). In addition, this trial will undertake comparative assessments between surgical intervention combined with CDT and CDT alone across various domains, encompassing changes in arm volume, frequency of lymphatic drainage, incidence of lymphangitic events, pain levels, further patient-reported outcomes (PROs) as well as economic considerations such as the reduction in the frequency of outpatient visits (secondary objectives). The trial will be conducted comparing parallel groups allocated in a 1:1 ratio.

## METHODS AND ANALYSIS

### Aims

Microsurgical approaches represent viable treatment options for patients suffering from chronic BCRL who do not achieve sufficient relief through CDT alone. Based on the current evidence available in the literature, microsurgical techniques are hypothesized to be superior to CDT alone.^(37–46, 48, 49)^

#### Primary Objective

To assess the effects of lymphatic surgery compared) combined with CDT to CDT alone for patients with chronic LE on long-term lymphedema-specific QoL (at 15 months) using the Lymph-ICF-UL questionnaire.

#### Secondary Objectives

- To test whether lymphatic surgery provides better QoL compared to CDT with respect to additional short- and long-term QoL outcomes and better patient satisfaction.
- To compare lymphatic surgery and CDT with respect to arm volume, frequency of lymphatic drainage and lymphangitic events.
- To compare the burden on patients of lymphatic surgery and CDT, in terms of total number of operative procedures, length of hospital stay and total number of outpatient visits.
- To compare lymphatic surgery and CDT with respect to economic aspects.
- To compare the safety of lymphatic surgeries LVA and VLNT, in terms of surgical complications.

### Design

This is a pragmatic, randomized, international, multicentre superiority trial comparing microsurgical treatment of BCRL combined with CDT to conservative treatment with CDT alone, conducted in two parallel groups.

The PRECIS-2 tool was employed in the design of this pragmatic trial (design scores: Eligibility 4; Recruitment 5; Setting 5; Organization 5; Flexibility delivery 5; Flexibility adherence 5; Follow-up 4; Primary outcome 5; Primary analysis 5).^(50)^

Randomization will be carried out utilizing a standard minimization algorithm to maintain balance between patients assigned to receive either of the two treatments (surgical vs. non-surgical treatment). This will result in a 1:1 allocation with a total of 140 patients in each treatment arm. Minimization will also achieve balance across study sites, state of LE and planned surgical technique.

The study duration is planned to span 13 years, including approximately two years for recruitment (starting in Q3/2023), primary endpoint analysis after 15 months of follow-up of the last patient and the end of the main study 24 months after last patient inclusion. The extended follow-up will conclude 10 years after the last patient inclusion (approximately Q3/2035) with the long-term analysis expected to be finalized approximately one year later (Q3/2036).

**Figure 1:**
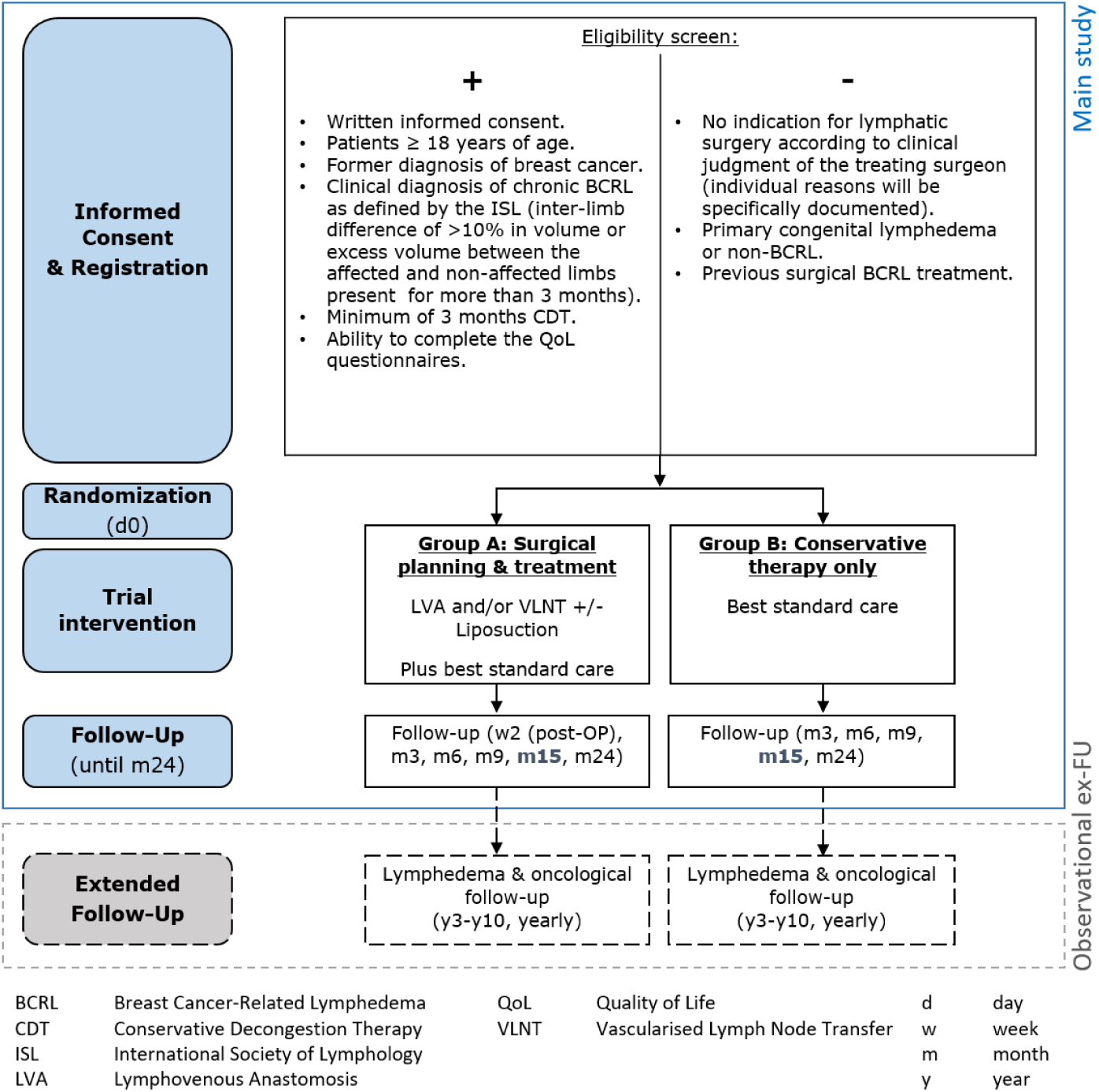
Flow chart of study design.

### Study setting

Over 20 international sites across Europe, USA, Canada and Latin America will participate in the study. The study sites consist of academic, public and private hospitals that regularly provide lymphatic surgery. The list of study sites can be obtained at lymphtrial.com. The study was initiated at the University Hospital Basel (Switzerland) in July 2023, and by February of the following year, the first 13 patients in Switzerland had been enrolled.

### Participants

Participants aged 18 years and above who have previously undergone treatment for breast cancer and have received a clinical diagnosis of chronic BCRL are be eligible for this trial. BRCL is defined in accordance with the definition established by the International Society of Lymphology (ISL), which involves an inter-limb volume difference exceeding 10% or an excess volume in the affected limb compared to the non-affected limb, persisting for a duration of more than three months. Patients must have undergone CDT for at least three months and possess the capability to complete the QoL questionnaires. This study will exclude patients with primary congenital LE, LE unrelated to breast cancer treatment and previous surgical BCRL treatment.

### Intervention and procedures

#### Group A: Surgical Group

According to the pragmatic study design, neither the diagnostic workup nor the surgery will be standardized in order to offer surgeons considerable leeway on how to perform lymphatic surgery, which resembles the flexibility in usual care. However, the key aspects of the preoperative workup and the surgery including the number of LVAs, harvesting of lymph nodes (“donor site”), time of surgery, and practical details will be registered.

LVA and VLNT are both surgeries with limited invasiveness to treat BCRL. However, to date it remains largely unclear, if one of the two techniques is superior compared to the other or if the severity of BCRL (stage of LE) favours one technique over the other. Nevertheless, it is not within the scope of this trial to compare one surgical technique to the other, but rather to examine whether physiological or reconstructive LE surgery, is superior to CDT alone.

##### LVA

The objective of this procedure is to create a lymphovenous bypass, to redirect lymph to the venous system without lymphatic drainage through the thoracic duct.^(28, 41, 51–54)^ Surgery is performed under general anaesthesia to avoid patient discomfort using a high-magnification microscope and specific super-microsurgical instruments and sutures, as lymphatic vessels can be less than 0.8 mm in diameter. The anastomosis is usually performed in an end-to-end fashion in case both the lymphatic vessel and the vein have approximately the same diameter, otherwise, in cases of mismatch of vessel diameter, an end-to-side anastomosis is preferred. The patency of the LVA is confirmed by direct visual examination under the microscope and/or intraoperative angiography using a specific contrast media (indo-cyanin green).

##### VLNT

The procedure is based on replacing lymph nodes (LN) that have been previously resected surgically and/or harmed by radiotherapy in the axilla, using a vascularized composite flap containing LNs (between three and six nodes), adipose tissue and in some cases skin, harvested from a specific donor site. Various different donor sites can be chosen in one patient,, such as axilla, groin, submental region, omentum.^(28, 38, 55–57)^

##### Liposuction

Depending on the surgeon’s preference, lympho-liposuction can be used in conjunction with LVA/ VLNT or as a separate “stand alone” procedure.^(58)^ Lympho-liposuction is often performed using vibrating cannulas that facilitate the process. It is mostly performed following the technique described by Brorson.^(59)^

##### Two-stage LE surgery

If applicable, LE surgery can be performed in two stages at the discretion of the treating physician, which will be documented. Traditionally LE surgery has often been performed as a two stage procedure, recent data though suggests that a one-stage approach is equally safe and entails several advantages, such as avoiding secondary interventions which increase the burden on both patients and on the healthcare system.^(60)^

##### Pre-surgical visit

According to local standard of care, pre-surgical outpatient visit(s) will be performed e.g. to discuss details of the surgery together with the patient, to obtain written consent for surgery or to perform lymphangiography for surgical planning.

##### Postoperative treatment

According to the pragmatic study design, the postoperative treatment regimen will not be standardized either. However, the key aspects like frequency and timing of lymphatic drainage, as well as class of compressive garments and the use of peri- and postoperative antibiotics will be registered. We recommend that immediately after the surgery, the patient’s arm is bandaged for compression. The day after surgery, CDT should be reinitiated excluding the stitched site(s). Shortly after the removal of surgical stitches (approximately 2 weeks post-surgery), we recommend that bandages will be replaced by the respective compressive garments.^(61, 62)^

#### Group B: CDT (Control group)

CDT will be performed as in usual care, following the pragmatic study design. The key aspects like frequency of lymphatic drainage, time when lymphatic drainage is performed and time and class of compressive garments are used will be documented.

CDT incorporates two stages of treatment. The first treatment phase (intensive phase) entails skincare, MLD, exercises aimed at improvement of mobility/range of motion in the shoulder, elbow or wrist joints, and compression therapy through bandaging. Most patients undergo this phase shortly after the diagnosis of LE. CDT in the second phase (maintenance phase) aims to maintain the achieved limb volume/ circumference reduction through compression with therapeutic elastic compression garment for the arm.^(26)^ Skincare, mobility exercises and MLD is continued in this phase if needed.^(26, 27)^

In order to reduce LE to the maximum with CDT, we recommend to intensify treatment without having more appointments. For this, the above described first and second phases should be alternated in a customised regime to sustainably reduce LE without losing patient adherence, with less time consuming and at the lowest possible cost. Therapy starts with an intensive phase lasting one to two weeks to reduce edema volume.^(63)^ Therapy contains manual lymphatic drainage in an evidence based manner^(64)^ and including intensive techniques as promoted by Belgrado.^(65)^ In this time wearing the bandaging with ca. 20mmHg pressure^(66)^ is mandatory for the time between appointments. At the end of this phase a custom made flat knitted stocking is issued. In the subsequent maintenance phase, self-therapy (wearing the stocking and exercises / sports) is carried out. It is not necessary to continue MLD once or twice every week in this phase.^(67)^

After the primary endpoint assessment (month 15) patients in the control group will be offered the possibility of a crossover to the surgery arm. In this case the same surgery visit schedule will be followed and the respective details documented. Further follow up visits of the surgery will follow the routine care, the study schedule e.g. 24 month visit after randomization.

### Withdrawal and discontinuation

Patients have the right to discontinue their participation in the trial for any reason and at any time, without prejudice to further treatment.

Once a patient is randomized, the study site will make every reasonable effort to follow the patient during the entire study period. The anticipated loss to follow-up rate is maximally 10%, which has been used to adjust sample size calculations. The time points for study visits have been matched with the schedule of standard clinical visits after surgery according to the pragmatic trial design.

If a patient withdraws her consent (i.e. refuse further data collection), she/he will be informed that all data collected until the time point of their withdrawal will be kept coded and used. For the patient’s security, a last examination will be performed.

### Adherence

Since in most cases the surgical intervention takes place only once, adherence is not a relevant issue in the surgery group. Patients in the control group without surgery will be encouraged to have no surgery throughout the two years after randomization, but due to the pragmatic nature of the study, no specific measures to increase adherence are taken. Patients without surgery will follow the same outpatient visit protocol as the patients in the surgical group and will be encouraged to continue CDT.

### Concomitant care

All relevant concomitant care and interventions are permitted and none are prohibited during the study thus reflecting the pragmatic nature of the trial. Concomitant therapy includes radiotherapy or systemic therapies such as chemotherapy, anti-hormonal therapy, immune checkpoint-inhibitor therapy, T-DM1 therapy or other relevant therapies such as rehab for intensive lymphatic drainage.

### Outcomes and assessments

#### Primary Endpoint

The primary endpoint of this trial is the patient reported QoL outcome “lymphedema-specific QoL”, which will be assessed 15 months after randomization (and therefore about 12 months after surgery) measured by the Lymph-ICF-UL questionnaire.

#### Secondary Endpoints

If not specified, secondary endpoints will be assessed at various time points according to the visit plan.

Safety Endpoints: Adverse events; complications of surgery *(applicable in surgery group only)*, lymphangitic events (erysipelas).

PROs: QoL-Lymph-ICF-UL; QoL-LYMPH-Q; QoL-EQ-5D-5L; Pain score (VAS).

Additional Endpoints: Lymphedema assessment including arm volume; frequency of lymphatic drainage; burden on patients (total number of operative procedures, length of hospital stay, absences from work and number of out-patient visits); economic evaluation (for Switzerland) including condition-related medical resource use, condition-related healthcare costs, condition-related indirect costs, quality-adjusted life-years and incremental cost-effectiveness.

#### Influence of baseline factors on the endpoints

Patient-dependent baseline factors that may have an impact on the endpoints are stage of LE and baseline Lymph-ICF-UL score.

### Detailed description of the assessments

#### PROs

Patients will be instructed to complete all relevant questionnaires (details described below), which will be completed by the patients at the beginning of each study visit. Only officially translated and validated PRO questionnaires will be used within this trial. Patients can only participate in the trial, when they are willing and able to complete the PRO questionnaires, e.g. being proficient in the available QoL languages.

Baseline questionnaires will be completed before randomization. For the subsequent assessments, the QoL questionnaires will be completed before any diagnostic procedures or communication of diagnostic or prognostic information to the patient, and before any treatment or supportive care measures.

The questionnaires are answered by the patients themselves, either on paper or if possible directly electronically in the CDMS.

##### Lymphedema-specific QoL - Lymph-ICF-UL

One of the most widely used PRO instruments in chronic BCRL is the LYMPH-ICF, a rigorously developed and validated PRO instrument specific to BCRL with various translations available. The LYMPH-ICF-UL has been used in thousands of patients.^(37, 51, 53, 68–77)^ This questionnaire assesses the impairments in function, activity limitations and participation restrictions of patients with upper limb LE. It is a validated questionnaire, consisting of 29 items (questions) across five different domains

##### Lymphedema-specific QoL - Lymph-Q

The Lymph-Q Upper Extremity Module is a new PROM developed to assess patient reported outcomes of BCRL in a concept-driven approach. The questionnaire was designed together with women treated for breast cancer who developed arm LE and then field-tested in 3222 women with arm LE from the USA and Denmark.^(78)^ The complete questionnaire contains 68 questions covering the patient-relevant topics health-related quality of life, experience of care, and treatment.

##### Pain score

The pain score used consists of a VAS ranging from 0 (i.e. no pain) to 10 (i.e. worst pain), published by the Yale University.^(79)^

##### Health Economics - EQ-5D

The EQ-5D is one of the most widely used instrument for measuring health-related QoL for cost-effectiveness analyses.^(80)^ The 5-level EQ-5D version (EQ-5D-5L) consists of the EQ-5D descriptive system that comprises five dimensions: mobility, self-care, usual activities, pain/discomfort and anxiety/depression and will be used for the trial.^(81)^

#### Safety outcomes

##### Surgical complications

Surgical complications will be assessed at each study visit (applicable in surgery group only) according to the modified classification of Clavien-Dindo.^(82)^

Surgical complications at the LE site(s) as well as at the lymph node donor site(s) will be assessed as follows:

- <u>Haematoma</u> - clinically relevant, defined as either causing discomfort, or requiring intervention
- <u>Seroma</u> - clinically relevant, defined as either causing discomfort, or requiring intervention
- <u>Wound infection</u> - treated with oral or intravenous antibiotics, with (major) or without (minor) surgical exploration
- <u>Wound dehiscence</u> - opening of an originally closed surgical wound due to various reasons like secretion, infection, reaction to suture, drains or insufficient tissue perfusion
- <u>Donor site LE</u> – LE (ISL grade ≥ 2) in the lower limb due to lymph node harvesting from this area for VLNT

##### Lymphangitic events (erysipelas)

Lymphangitic events are defined as skin infections at the LE site(s) which can be treated with oral or intravenous antibiotics.

#### Oncological Outcomes

To address the oncological outcome of patients, recurrence-free survival (RFS) will be determined. RFS is defined as the time from oncological surgery until the first documentation of any of the following events: local-regional occurrence or recurrence of invasive disease or ductal carcinoma in situ (DCIS), distant breast cancer metastasis, or death from any cause.

#### Additional outcomes

##### LE Assessment

At baseline and primary endpoint visit, the stage of LE will be classified according to the ISL. The excess limb volume is measured as the difference in volume between the affected and unaffected limb which is reported as a percentage of the volume of the unaffected limb.^(83, 84)^ A relative volume reduction well as an absolute volume reduction will be calculated.^(85)^ Arm circumference will be compared to the healthy contralateral arm. As there is no standard number of measurements per arm, the assessment in this trial referred to existing guidelines: the National LE Network advises six circumference measurements at minimum^(86)^ and the ISL recommends that measurements be taken at 4cm intervals^(87)^. Striking a balance between the two guidelines and aiming to lessen the load on patients, the arm circumferences will be measured in 10 cm intervals from the wrist of the hand at each visit ^(87–89)^ These are routine measurements, done by measuring tape by the treating physiotherapist on regular consultations. To ensure measurements at the respective visits, all LE assessments will be performed by trained study personnel.

Other data collected within LE assessment include arm aesthetics assessments by patient and physician, photographic documentation of both arms (at baseline and after 15 months), physical exam of skin and axilla, and arm shoulder motion.

##### Conservative therapy and lymphatic drainage

Details on conservative therapy and the frequency of lymphatic drainage will be registered at the baseline visit as well as at each follow-up visit. General collected information include frequency of therapy sessions before the start of the study and between the study’s follow-up visits, time frame and type of compression garments worn, and performance of MLD and skin care. Additional, patient’s treating physiotherapists will be asked to record routinely assessed measurements, such as arm volume measurements, tissue quality assessments and shoulder motion test.

##### ICG mapping or other imaging methods

Depending on the centre, various imaging methods will be performed at the screening or preoperative visit in different centres following the local standard and competence. The different methods/results will be assessed in detail to allow for later comparability between methods. ICG mapping will be graded according to the MD Anderson grading scale ^(61, 90)^ and Arm dermal backflow scale ^(91)^.

##### Burden on patients

The burden on patients will be assessed by different means: Total number of breast-/arm-related operative procedures; length of LE-related hospital stay(s); total number of LE-related outpatient visits; LE-related change in the ability to work and number of missed working days and hours.

#### Baseline variables

Baseline variables will be assessed at the screening visit and include patients’ demographics & characteristics, general wellbeing (physician assessed Karnofsky performance score^(92)^), personal and medical history, previous treatment, previous surgery of the breast/axilla, previous oncological therapy, previous postmastectomy radiotherapy and tumor characteristics.

#### Magnetic Resonance Lymphangiography (MRL) and Magnetic Resonance Imaging (MRI)

The additional MRL and MRI acquisition and evaluation will be performed only for a subgroup of patients enrolled at University Hospital Basel or if it is standard of care in other study sites. LE grading as well as skin and lymphatic channel assessment will be performed by a plastic surgeon and a radiologist jointly and independently from the ICG grading. The concordance (correlation) between MRL and ICG coordinates for lymphatic vessels will be performed as in Pons G. et al.^(93)^

#### Economic assessments (from a Swiss perspective)

The following parameters will be assessed within the first 24 months after randomization as well as for the extended FU time: Condition-related medical resource use; condition-related healthcare costs, based on, in- and outpatient costs accrued at the treating site as well as with other healthcare providers; condition-related productivity losses; QALYs, incremental cost-effectiveness.

### Participant timelines and procedures at each visit

Study duration of each patient is 10 years. The first 2 years are the interventional part including the primary endpoint assessment after 15 months and end of main study follow-up (after 24 months). Afterwards the patients will enter the observational part for another 8 years of follow-up.

#### SCR(Screening)/Baseline/Enrollment (up to -30 days before V1)

Prior to study registration the following steps have to be performed: Informed consent of the patients obtained latest at the Screening Visit (SCR) (but can be obtained earlier, up to 6 months before V1), baseline variables assessed, patient listed in the Screening and Enrollment log, eligibility criteria for registration checked. Patients can then be registered via the web-based CDMS secuTrial®.

Procedures: Screening, Informed Consent, Registration, Baseline variables, LE assessment, Photographic documentation of the arms, Physical examination of skin and axilla, ICG mapping or other imaging methods, Conservative therapy/ lymphatic drainage, Lymph-ICF-UL, Lymph-Q, EQ-5D-5L, Pain score, Burden on patients

#### V1/Randomization/d0

Patients will be randomized to either CDT or surgical treatment group. Patients randomized to the control arm will receive the standard of care treatment CDT. A treatment example/suggestion is described in detail above, but according to the pragmatic study design, CDT will not be standardized. Patients randomized to the interventional arm should undergo surgical treatment (LVA or VLNT with or without liposuction) as soon as possible but latest 3 months after randomization. Patients randomized to the interventional arm will also receive CDT.

##### Procedure: Randomization

###### V1-1/Pre-surgical planning (Only for patients receiving surgery)

A pre-surgical planning visit can be performed according to the local standard of care. If not performed yet, ICG mapping or other imaging methods used in the local routine care for surgical planning can be performed during the visit. A subgroup consisting of patients at the University Hospital Basel, Switzerland will also undergo magnetic resonance lymphography (MRL) and magnestic resonance imaging (MRI) in addition to ICG mapping irrespective of the randomized study intervention group.

Procedures: ICG mapping or other imaging methods (*Only if this is local standard of care at any site)*, MRL and MRI (*Only if this is local standard of care (any site) or if the patient is included at the site in Basel*

###### V1-2/Surgery (< 3 months after V1) (Only for patients receiving surgery)

Patients will be operated according to the local standard of care.

Procedures: Intervention, Surgical complications, Lymphangitic events, (Serious) Adverse events

###### V1-3/Post-surgery/2 weeks (+/- 7 days) after V1-2 (Only for patients receiving surgery)

Patients who received surgery will have an additional follow-up visit two weeks after the surgery appointment according to the routine care.

Procedures : LE assessment, Lymph-ICF-UL, Lymph-Q, EQ-5D-5L, Pain score, (Serious) Adverse events, Burden on patients, Concomitant care

Follow Up: *V2/month3 (+/- 14 days), V3/month6 (+/- 1 month), V4/month 9 (+/- 1 month), V5/month15 (+/- 1 month), V6/month24 (+/- 1 month)*

Follow-up visits will be performed at month 3, 6, 9, 15 and 24 after randomization. Procedures: LE assessment, MRL/MRI *(at V2 only, only if not done before (SCR, V1-1)* and o*nly if this is local standard of or in Basel)*, Photographic documentation of the arms (*only at V5)*, Conservative therapy/ lymphatic drainage, Lymph-ICF-UL, Lymph-Q, EQ-5D-5L, Pain score, Surgical complications (*only for patients receiving surgery*), (Serious) Adverse events, Lymphangitic events, Concomitant care, Burden on patients, Oncological outcomes *(only at V4 – V6*), Cost data from hospital administrations (*only at V6 and only for patients included at Swiss sites*)

###### Extended FU (exFU 1-8) (from 3 to 10 years, yearly (+/- 2 months))

In order to be able to determine long-term outcomes of lymphatic surgery, an observational yearly follow-up will take place after each patient has completed the 24 month from 3 years to 10 years after randomization. Patients will be invited for a consultation visit in the hospital. In the case of excessive effort (e.g. too long travel time), a remote visit can be performed via a telephone call. QoL questionnaires will in this case be sent to and filled out by the patient at home.

Procedures: LE assessment, Conservative therapy/lymphatic drainage, Lymph-ICF-UL, Lymph-Q, EQ-5D-5L, Oncological outcomes, Burden on patients

### Sample size

The sample size was calculated with the objective of detecting a difference in the primary endpoint (LE-related QoL at 15 months) between the two study groups, at a significance level α = 5%.

The clinically relevant difference in the Lymph-ICF-UL score between surgical techniques and CDT was defined as θ = 10 points on a scale ranging from 0 to 10. This choice was made in consultations with the patient advocacy group, as an improvement of 10 points in the total Lymph-ICF-UL score corresponds to altering three responses from “not at all” to “very well”, or enhancing each individual response by one point. The choice of a 10 point difference as being clinically relevant is supported by the data of Devoogdt et al. and De Vrieze et al. ^(94, 95)^, as it aligns with 2 standard deviations of the within-subject variability in the total Lymph-ICF-UL score. Moreover, the findings of Qiu et al. ^(51)^ demonstrate that achieving such an improvement in the primary endpoint is clearly feasible.

Based on the results documented by Devoogdt et al. and De Vrieze et al.^(94, 95)^, it was assumed that the Lymph-ICF-UL scores for both surgical techniques and CDT follow an approximately normal distribution with a standard deviation of σ = 21.9. A Student’s t-test will be used to compare the average Lymph-ICF-UL scores between the two study groups.

With an anticipated drop-out rate of 10% and a total arm-switching rate of 5% (corresponding to 10% of patients in the CDT arm switching to the surgery arm and no patients in the surgery arm switching to the CDT arm), the recruitment goal is set at 280 patients. This number is intended to yield a total of N = 252 evaluable patients (126 in each study arm), providing a power of 90% when the absolute treatment effect is θ = 10.

### Recruitment plan

Patients will be recruited by reconstructive surgeons, specialized in the (micro) surgical treatment of LE. Enrolment will take place at the outpatient clinics of the participating hospitals. According to the accrual estimates, over 20 sites are adequate to recruit the required 280 patients during the 24-month recruitment period. To ensure a good recruitment rate, more sites than theoretically needed will be asked for trial participation and opened as soon as possible. Two back-up strategies are pre-specified in case of under-recruitment. First, estimated versus actual accrual will be continuously monitored for each study site with an early first evaluation of recruitment already six months after opening the first site. If the trial is under-recruiting, the second back-up strategy includes further international escalation, e.g. inclusion of additional backup study sites.

### Assignment of intervention

#### Randomization

The randomization process will be implemented within secuTrial®, the clinical data management system. Randomization will be executed using a standard minimization algorithm to achieve equal distribution of patients between the two treatments (surgical vs. non-surgical treatment) in a 1:1 ratio totaling 140 patients per treatment arm. The minimization will balance the study site, the stage of the patient’s LE and the planned surgical technique. To avoid a predictable alternation in treatment allocation, patients will be assigned to the treatment group that minimizes the imbalance between the two treatment groups within each study site, with a probability of 80 percent, without the need for a allocation list/ sequence.

Every patient who consents to participate and meets the inclusion criteria will be randomized. The investigators or delegated study personnel will enroll the patients. The randomization occurs during or after the screening visit a maximum of three months prior to the potential surgery date. The investigator or their delegated study personnel informs the patient about the randomization outcome either directly in person or over the phone and schedules the next appointment accordingly.

#### Blinding

Blinding the patients, the study team or other caregivers is not feasible due to the nature of the proposed surgeries (LVA and/or VLNT with or without liposuction) resulting in visible scars within the surgical group. Moreover, the primary outcome and some of the secondary outcomes are patient-reported outcome measures, which are inherently subjective in nature. Objective measurements for certain secondary outcomes (e.g. volume measurement), cannot be blinded and will be conducted by trained personnel at the study site. Both the surgical team and the physiotherapists will adhere to established clinical standards regarding the diagnosis as well as the treatment of complications. Given that these professionals primarily responsible for complication management are not blinded, un-blinding procedures will not be required.

## METHODS AND ANALYSES

### Data collection methods

Investigators and other study personnel at each center will receive centralized training on the study’s specific requirements. This training will include a comprehensive review of the data to be collected and the procedures to be carried out. In addition, investigators’ meetings will provide a platform for discussing the details of data collection forms and the type of information required. The investigators and study staff will also receive instruction on how to utilize the CDMS.

All questionnaires will be validated in the languages used and will be provided to the study sites. Patients can complete the questinnaires on paper or if possible directly electronically in the CDMS. In addition worksheets and checklists for the assessments will be provided to the sites along with clear and comprehensible instructions, to ensure data integrity. Data entry will be performed by trained clinical investigators and trained study personnel.The principal investigator at the study site is responsible that the data entered into the eCRF are complete and accurate, and that the entry and updates are performed in timely manner.

#### Retention

To ensure ongoing engagement of all enrolled participants, we strive to sustain their interest in the study by actively maintaining our website, scheduling appointments early and promptly addressing any issues or inquiries they may have. The causes behind non-compliance and participant attrition will be recorded. All participants will be included in an intention-to-treat analysis, irrespective of their adherence.

### Data management

Data will be recorded both in hard copy format and electronically within the CDMS. If study worksheets/ paper forms are used, they will be securely stored at the respective participating site and the data will be transferred into the CDMS by the sites. Study data will be captured via the online CDMS secuTrial®, based at the IT-department of the University Hospital Basel. The CDMS is accessible via a standard browser on devices with internet connection. Password protection and user-right management ensures that only authorized study personell has access to the data during and after the study. Backup of secuTrial® study data is performed regularly according to the processes of the IT department of the University Hospital Basel. Study data entered into the eCRF is only accessible by authorized persons.

An audit trail will maintain a record of initial entries and any changes made; time and date of entry; and username of person authorizing entry or change. For each patient enrolled an eCRF must be completed. A unique patient identifier will be used to identify patients.

The data will be reviewed by the responsible investigator as well as an independent monitor. The monitor will raise queries using the query management system implemented in secuTrial®. Designated investigators have to respond to the query and confirm or correct the corresponding data. Thereafter the monitor can close the query.

### Statstical methods

#### Statistical analysis plan

Detailed methodology for summaries and statistical analyses of the data collected in this study will be documented in a statistical analysis plan. The statistical analysis plan will be finalized before database closure and will be under version control at the Department of Clinical Research, University of Basel and University Hospital Basel.

If substantial deviations of the analysis as outlined in these sections are needed for whatever reason, the protocol will be amended. All deviations of the analysis from the protocol or from the detailed analysis plan will be listed and justified in a separate section of the final statistical report.

#### Planned analysis

All analyses will be conducted using the statistical software package R^34^, using “two-sided” statistical tests and confidence intervals with standard significance and confidence levels α = 5% and (100 % – α) = 95%, respectively.

The null-hypotheses are that there is no difference between the two study arms (i.e. no treatment effect) as regards the primary and secondary endpoints. The alternative hypotheses are that there are differences between the two study arms.

The full analysis set (FAS) will include all patients who were randomized. The per protocol set (PPS) will include all patients within the FAS who fulfilled the eligibility criteria and for whom the treatment and follow-up were completed as planned in the study protocol. All statistical analyses will be performed on the FAS according to the intention-to-treat principle (i.e. all patients will be analyzed on the basis of the study arm to which they were randomly allocated), except for sensitivity analyses performed on the PPS.

Baseline characteristics of all patients in the FAS and PPS will be summarized, ungrouped as well as grouped by study arm (missing values will be ignored, but the proportion of missing values will be reported for each variable). Categorical data will be presented as absolute and relative frequencies, while for each numerical variable, the mean and standard deviation, or the median and interquartile range will be presented, as appropriate. The standardized mean difference between study arms will be reported for each baseline characteristic.

The treatment effect on the primary endpoint will be examined by analysis of covariance (ANCOVA), adjusted for baseline Lymph-ICF-UL score and LE stage. Several sensitivity analyses will be performed: on the PPS, without covariates, with interaction between treatment and LE stage, and with study center as an additional covariate (with and without interaction with treatment).

The treatment effects on the secondary endpoints will be examined by ANCOVA or appropriate generalized linear models, adjusted for baseline value and LE stage (with and without interaction with treatment). The primary and secondary analyses will be repeated with surgical treatment (VLNT, LVA, liposuction) as an additional covariate (with and without interaction with treatment).

Safety will be assessed via a rigorous and detailed examination of adverse events, serious adverse events, complications of surgery and lymphangitic events.

#### Additional analyses: translational research

Multiple subprojects encompass a range of open questions related to oncology, therapeutics, economics and surgeries.

- Comparison of LE specific QoL
- Impact of timing of surgery on outcome
- Impact of stage of LE on surgical outcome
- Patient referral: Analysis which type of physicians refer patients for lymphatic surgery.
- Health economic evaluation
- Role of (MRL/MRI) imaging on surgical decision making
- Effect of radiotherapy on microsurgical BCRL treatment and LE outcome
- Correlation of breast cancer radiation dose on stage of LE and lymphatic surgery outcome.
- Impact of number of LVAs on surgical outcome
- Impact of localization of LVA on surgical outcome
- Impact of number of transferred lymph nodes on surgical outcome
- Correlation of patient-reported arm aesthetics, QoL and physician’s-reported arm aesthetics
- Impact of axillary scar release on outcomes of VLNT surgery
- Impact of patient and public involvement on clinical trial success: evaluation of factors determining successful patient inclusion into the study and its effect on trial outcomes
- Comparison of surgical complications between LVA and VLNT
- Assessment of crossovers from the CDT to the LE surgery arm
- Correlation between localization of breast cancer and stage of LE as well as outcome of LE surgery
- Effect of chemotherapy (and timepoint of chemotherapy) on LE and microsurgical BCRL treatment
- Analysis of the impact of surgery or surgical technique (LVA or VLNT) on arm aesthetics
- Comparison of LE stage between baseline, primary endpoint and extended follow-up
- Comparison of the arm volume measurements between trained study site personnel and treating physiotherapists
- Assessment of seasonal effects on QoL
- Analysis of the influence of (simultaneous) breast surgery/reconstruction type on development of LE
- Impact of experience of study site on outcome of LE surgery (Lindenblatt?)
- Impact of additional liposuction vs. reconstructive surgery only (VLNTx, LVA) on outcomes (PROMs and objective).
- Effect of surgery on arm measurements by number of lymph nodes removed during index axillary surgery
- Effect of surgery on primary quality of life endpoint by history of psychiatric diagnosis (anxiety and/or depression)
- Trends in use of LVA vs VLNT by site and by country
- Performance of LVA vs VLNT
- Performance of microsurgical lymphatic surgery with vs. without Biobridge®

The detailled statistical analyses of the subprojetcs will be described in the statistical analysis plan, which will finalized before database closure.

#### Handling of missing data and drop-outs

Missing values will be handled by available-case analyses. However, if such analyses exclude more than 5% of the patients, sensitivity analyses with multiple imputation by chained equations based on the MAR (missing at random) assumption will be considered.^(96)^ A drop-out rate of 10% was taken into account in the sample size determination.

After 90% of the required patients (= 252 patients) have been recruited, both the number of patients who dropped out and the number of patients who did an early emergency cross over will be reviewed in a blinded interim analysis. If the drop-out or crossover rate is higher than estimated, it can be decided that additional patients can will be recruited to reach the final number of patients for meeting the primary endpoint.

### Monitoring

#### Data monitoring

The study follows a risk-adapted monitoring approach, which is comprehensively described in the study monitoring plan. Oversight of the trial’s safety will be conducted by an independent data safety monitoring board (DSMB), comprising a study-independent statistician and three independent experts with expertise in chronic BCRL, LE surgery and clinical trials. The DSMB will evaluate the safety of the trial and suggest appropriate measures if necessary. The board may recommend holding the trial should serious complications arise in either group. The steering committee will generally follow the recommendations of the DSMB.

#### Harms

Adverse events (AEs) of interest, which are surgical complications, lymphangitic events and oncological outcomes, will be documented during each study visit, as well as during the extended follow-up. Surgical complications will be assessed in the surgery group only. Patients will be instructed by the investigator to report the occurrence of all AEs. Lymphangitic events at the LE site(s) will be documented for all patients. To assess the oncological outcomes, recurrence-free survival (RFS) will be recorded.

Serious adverse events (SAEs) associated with the compared interventions in both groups (surgery and conservative treatment) will be documented and reported to the sponsor– investigator within 24 hours. The Sponsor ensures that the event is reported to the respective ethics committee (EC) and/or authorities according to the national regulations. SAEs will be documented and reported only if there exists an assumed plausible association (possibly, probably, definitely) between the event and the interventions. Adverse events associated with breast cancer (treatment), breast cancer surgery or planned hospitalizations (e.g. for second-stage surgery) are not classified as SAE. Consequently, they are exempt from the requirement of expedited reporting. If it cannot be excluded that the SAE may be connected to the intervention under investigation, the SAE is reported to the respective EC and/or authorities according to national regulations.

All AEs and SAEs will be monitored until they have subsided or until a stable condition has been achieved. Depending on the nature of the event, further follow-up may require additional tests, medical procedures as warranted and/or referral to either a general physician or a specialist.

If immediate safety and protective measures have to be taken during the conduct of the study, the local PI notifies the Sponsor of these measures within 24 hours. For immediate safety and protective measures in Switzerland the Sponsor will notify the responsible EC of these measures and of the circumstances necessitating them within 7 days. For sites outside Switzerland the Sponsor will report safety and protective measures to the responsible EC if applicable according to their national law. An annual safety report (ASR) is submitted once a year to the local EC (if applicable) by the Sponsor.

#### Auditing

For quality assurance the Sponsor via an independent trial monitor (audit), the EC, or authorities (inspection) may visit the study sites. Direct access to the source data and all study related files is granted on such occasions. All involved parties keep the participant data strictly confidential.

## ETHICS AND DISSEMINATION

### Ethics

This study is conducted in compliance with the protocol, the current version of the Declaration of Helsinki, the ICH-GCP, the HRA as well as other locally relevant legal and regulatory requirements.^(97–99)^

Prior to initiating the trial, the trial protocol, informed consent document and any other relevant documents will be submitted to the respective independent Ethics Committees (EC). The EC ‘Ethikkommission Nordwest-und Zentralschweiz has granted ethical approval for the lead investigator’s site (2023–00733, 22.05.2023).

Significant modifications to the study setup, study organization, the protocol and relevant study documents will be submitted to the EC and/or authorities for approval before implementation. After the approval significant modifications will be communicated to the local principal investigators, all investigators and trial site staff, clinical monitors, data safety monitoring boards, clinical trial registries (for significant amendments) and participants (if the specifications impacts the treatments and risks or other aspects that could lead to requiring an updated informed consent). Any deviations from the study protocol will be fully documented using the study specific protocol deviation form.

Before being admitted to the clinical trial, all participants must provide written consent to participate after receiving a clear explanation of the nature, scope, and potential consequences of the trial by the investigators or their designees, presented in a format understandable to them. Additionally, patients will be asked in a second consent if their coded trial data can be used for further research projects in the future, that have been approved by a respective EC. There are no anticipated harm and compensation for participants.

### Confidentiality

Trial and participant data will be handled with uttermost discretion and is only accessible to authorised personnel who require the data to fulfil their duties within the scope of the study. The sites will retain all essential documents according to ICH GCP. This includes copies of the patient trial records, which are considered as source data, patient informed consent statement, and all other information collected during the trial. These documents will be stored for at least 10 years after the termination of the trial.

On the CRFs and other study specific documents, participants are only identified by a unique participant number; therefore, coded non-genetic data will be used for the trial analysis. Identification of patients must be guaranteed at the sites. For this purpose, sites are requested to use the patient identification list specifically produced for the trial. These documents will safely be stored at the sites. Patient confidentiality will be maintained according to applicable legislation. Patients must be informed and agree to data transfer and handling in accordance with Swiss data protection law and respective local laws at the sites. The patient data will entered by the sites into the eCRF/CDMS secuTrial®.

Once all data is entered into the CDMS and monitoring is completed, the database will be locked and closed for further data entry. The complete dataset is then exported and transferred to the trial statistician and the Sponsor through a secure channel.

### Access to data

Metadata describing the type, size and content of the datasets will be shared along with the study protocol and case report forms on public repositories adhering to the FAIR principles. The DKF of the University of Basel will act as an independent Data Access Committee (DAC) and store the data at time of publication on secure servers, maintained and backed-up by the IT-department of the University Hospital Basel. Researchers who wish to reuse data may submit a project synopsis at dkf.unibas.ch/contact.

### Patient and public involvement

Patients have played a fundamental role in shaping the study’s research question and primary endpoints, which were determined by the patients’ input to closely align with patient needs. Their engagement persists throughout all trial phases. Patient advocates were actively involved throughout the protocol development process including the evaluation of the visit schedule and trial assessments, with three of them serving as a co-author of this manuscript. Furthermore the patient information and consent form was reviewed by a patient representative to ensure comprehensibility. Patient advocates further hold an important role in the running trial with a voting member in the trial steering committee, during which they e.g. assess site and patient feedback, engage in discussions regarding necessary protocol and

patient information updates, and proactively address recruitment and retention challenges. Their roles also extend to ensuring the accessibility and comprehensibility of trial results for all patient groups, including relevant patient communities through diverse communication channels, such as study publications, newsletters, webpages, and social media reports.

### Dissemination

The study results will be published in a peer-reviewed medical journal adhering to the Consolidated Standards of Reporting Trials (CONSORT) standards for RCTs and in accordance with good publication practice, regardless of the outcome.^(100, 101)^ Authorship for future trial publications will be determined based on the contributions made by authors. Metadata detailing the dataset’s type, size and content will be made available alongside the study protocol and case report forms on public repositories, in accordance with the FAIR principles (Findability, Accessibility, Interoperability and Reuse). An annual safety report is submitted according to the national regulations to the local ECs by the Sponsor-Investigator.

## Data Availability

Metadata describing the type, size and content of the datasets will be shared along with the study protocol and case report forms on public repositories adhering to the FAIR principles. Researchers who wish to reuse data may submit a project synopsis at dkf.unibas.ch/contact.

## ETHICS STATEMENTS

**Patient consent for publication:** Not required.

## ACKNOWLEDGMENTS

Our thanks go to the patients who participated in the development of the trial. We extend our gratitude to the generous funding provided by SNF, Rising Tide, and Krebsforschung. We thank the participating sites and dedicated principal investigators, whose substantial contributions have been essential in shaping the protocol and facilitating the study. In particular, we would like to acknowledge Prof. Christ Crain for her role in supporting and advancing the study through her mentorship and expertise in clinical research.

## FOOTNOTES

EAK, YH and AS are joint first authors.

KS, YHA and WPW are joint senior authors.

### Contributors

EAK, AS, FP, TMH, JS, MK, RM, MC, KR, A-LE, MS, YS, CG, LGH, ALP, YHA and WPW contributed to the study protocol. YH wrote the first draft and finalised the protocol manuscript by incorporating feedback from all authors. All authors critically revised the manuscript, provided essential input and approved the final manuscript.

### Competing interests

NL: Scientific advisor and consultant for Medical Microinstruments (MMI) Other authors: None declared.

**Patient and public involvement:** Patients and/or the public were involved in the design, conduct, reporting and dissemination plans of this study. Refer to the Methods & Analysis section for further details.

**Provenance and peer review:** Not commissioned; peer reviewed prior to submission.

**Biological samples:** There will be no biological samples collected.

